# Real-world smartphone data can trace the behavioural impact of epilepsy: A Case study

**DOI:** 10.1101/2024.01.01.23300494

**Authors:** Arthur R van Nieuw Amerongen, Anne Marthe Meppelink, Arko Ghosh, Roland D Thijs

## Abstract

Neurobehavioural comorbidities have a detrimental effect on the quality of life of people with epilepsy, yet tracking their impact is challenging as behaviour may vary with seizures and anti-seizure medication side effects. Smartphones have the potential to monitor day-to-day neurobehavioural patterns objectively. We present the case of a man in his late twenties with refractory focal epilepsy in whom we ascertained the effects of ASMs withdrawal and a focal-to-bilateral tonic-clonic convulsion on his touchscreen interactions. Using a dedicated app, we recorded over 185 days, the timestamps of 718,357 interactions. We divided the various smartphone behaviours according to the next-interval dynamics of the interactions by using a joint interval distribution (JID). We analysed JIDs during two ASM load transitions: before versus during tapering and restarting medication versus tapering. Cluster-based permutation testing revealed significant differences, with accelerated next-interval dynamics during tapering and a reversal upon medication restart. We also compared the JID of the fbTCS day to the average of the three days before, showing markedly slower next-interval dynamics on the day of the convulsion. This suggests that smartphone interactions’ temporal dynamics may help monitor neurobehavioural comorbidities in epilepsy.

## Introduction

Neurobehavioural comorbidities have a detrimental effect on the quality of life in people with epilepsy.^1,2^ Their impact on day-to-day behaviour is, however, difficult to monitor as the burden may fluctuate over time due to medication changes or seizure occurrence. The usual approach to quantifying these impairments is to use standardised tests and questionnaires in a hospital-based setting, thus only providing a snapshot in time.

Smartphones have the potential to monitor day-to-day neurobehavioral patterns objectively. Recent work demonstrated its value in various neurological conditions, including Parkinson’s disease and multiple sclerosis.^3,4^ We demonstrated that smartphone touchscreen interactions show increased behavioural ageing in people with epilepsy.^5^ Interestingly, we found a 10-year gap between the smartphone-based estimated age and the chronological age, which parallels findings in MRI studies.^6^ We postulated that, apart from neurodegeneration, drug-induced central nervous system side effects and the occurrence of seizures contribute to accelerated behavioural ageing in epilepsy.

We present a single case study of a man in his late twenties in whom we examined the effect of anti-seizure medication changes and seizures on smartphone touchscreen interactions. He participated in a prospective cohort study relating smartphone interactions to self-reported seizures for six months (NCT04617418). During the study, he was admitted to an epilepsy monitoring unit (EMU), for a five-day pre-surgical evaluation. He had refractory right temporal lobe epilepsy with weekly focal impaired awareness seizures and yearly focal to bilateral tonic-clonic convulsions. No aetiology was identified despite having had several MRI scans.

In this case, we leverage the ground truth information on seizures provided by the video-EEG recordings and the anti-seizure medication (ASM) withdrawal linked to the admission.

## Materials & methods

The Leiden-Den Haag-Delft Medical Ethics Committee approved the study, and the subject consented to participate. He installed the TapCounter app (QuantActions, Lausanne) on his smartphone, which recorded the timestamps of all touchscreen interactions and kept a seizure diary for six months. The data from the app was automatically transferred to a cloud-based server (taps.ai, QuantActions, Lausanne).

The subject participated in the study from May to November 2021. Seven hundred eighteen thousand three hundred fifty-seven touchscreen interactions were recorded over 185 consecutive days, with a median of 3,311 daily interactions (interquartile range (IQR) 2,531 to 4,766 interactions). He used lamotrigine and perampanel, which were reduced before admission and completely withdrawn during admission (figure 1a). He reported a total of 14 focal impaired awareness seizures outside the admission period. The five-day video-EEG recording documented nine focal aware seizures (FAS) with autonomic onset on the first day, four on the second day, one electrographic seizure whilst asleep on each of the first two nights, and one focal to bilateral tonic-clonic convulsion originating from the temporal lobe soon after wake-up on the fourth day.

**Figure 1.**
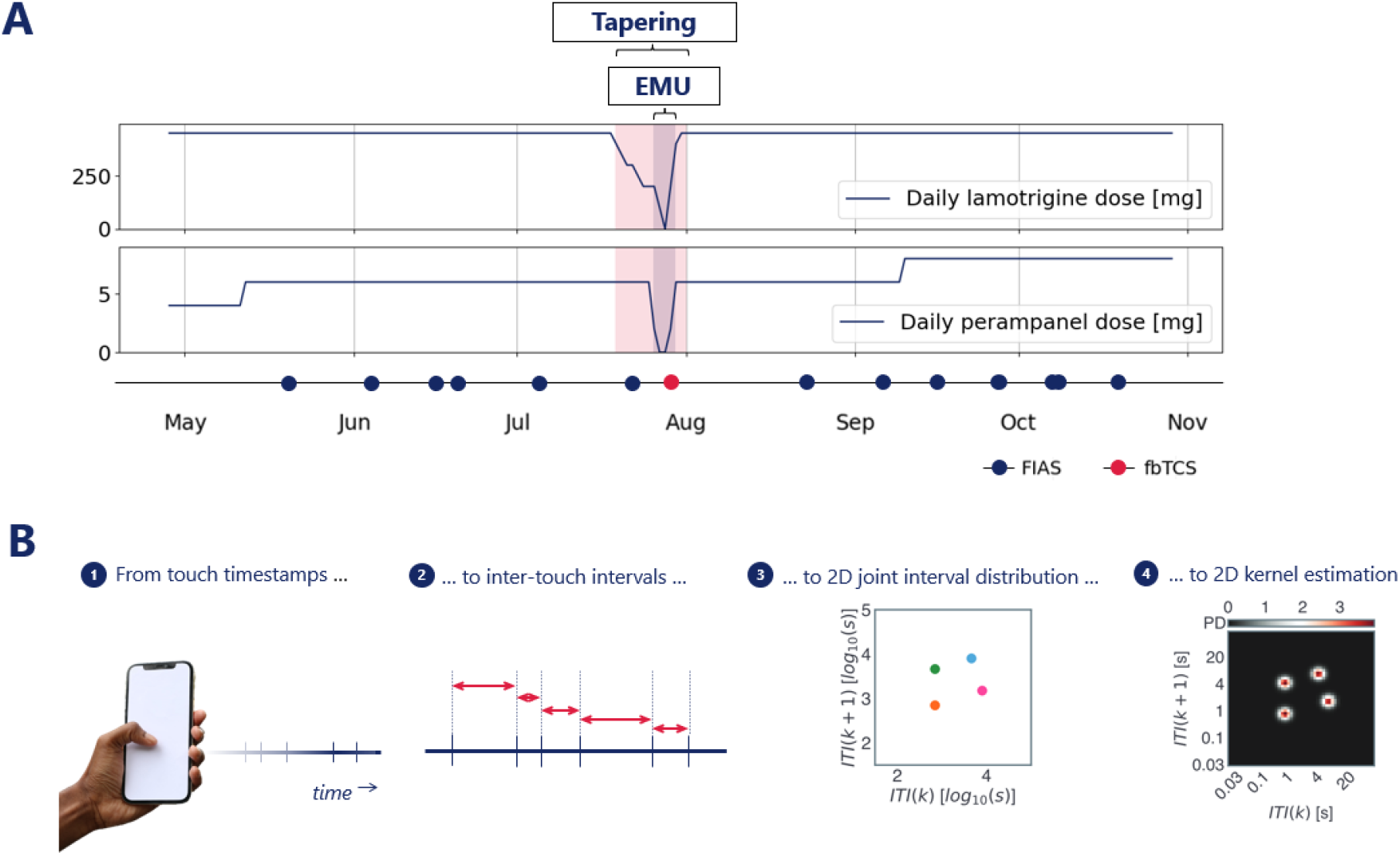
**A)** Timeline of the study period. The subject participated from May to November 2021. He reported several seizures, including oneconvulsion during admission. The anti-seizure medication load was stable for the study period; his medication was tapered before admission. **B)** A smartphone app recorded the timestamps of all touchscreen interactions (1). We calculated the inter-touch intervals (ITI) (2) and plotted each ITI at time k against the following ITI at time k+1 (3). We then estimated a 2D kernel density (4). **Abbreviations:** EMU: epilepsy monitoring unit; fbTCS: focal to bilateral tonic clonic seizure; FIAS: focal impaired awareness seizure; ITI: inter-touch interval

We downloaded the data from the online data collection manager (taps.ai, QuantActions, Lausanne). We applied a previously validated sleep and rise time detection algorithm to separate the touchscreen interactions per study day.^7^ We then calculated joint-interval distributions (JID) for the inter-touch intervals (ITI) from each day (figure 1b). We created a 2D space by relating each ITI (at time *k*) against its subsequent interval (at time *k + 1*). We operated 2D kernel density estimation (Gaussian kernel with a bandwidth of 0.1) over the log_10_-transformed 2D space. We subsequently discretised the continuous output of the kernel density estimation using 50 bins per dimension from ∼30 milliseconds to 100 seconds, thus obtaining a 50 by 50 matrix of 2D bins.^8^

To evaluate the impact of ASM tapering on touchscreen interactions, we categorised the data into three distinct periods: days before the ASM tapering (n=82), the tapering period itself (12 days), and days after ASM tapering (n=91). We evaluated the contrasts between the daily JIDs from the pre-tapering period vs. the tapering period, and the daily JIDs from the tapering period vs. the post-tapering period.

We employed a permutation-based clustering approach to identify clusters of 2D bins with significant differences in touchscreen interactions and to account for multiple testing associated with many individual t-tests (2500 t-statistics) when comparing JIDs bin-wise.^9^ Specifically, we performed 1000 random label permutations for each comparison (pre-tapering vs. tapering and tapering vs. post-tapering). At each iteration, we did a t-test for all 2D bins within the JIDs and identified clusters in the 2D space of t-statistics. We defined clusters as contiguous sets of 2D bins, where each bin has a t-statistic corresponding to a p-value of 0.001 or lower.

This method provided us with a reference distribution of maximal cluster sizes under the null hypothesis that both groups are indistinguishable. This allowed us to assess the statistical significance of cluster sizes observed in our original data. We considered a cluster size above the 95^th^ percentile of the reference distribution as statistically significant.

To assess the effect of seizures on touchscreen interactions, we compared the day of the convulsion during EMU admission (day 4) with the three days before. We first isolated the touchscreen interactions on the day of the convulsion and computed a JID for that day. Notably, this convulsion-related JID contains mostly post-ictal data, as the convulsion occurred shortly after awakening. We then computed the average JID from the three days before the convulsion and contrasted this distribution with that from the convulsion day to estimate the difference per bin in both JIDs.

## Results

The cluster-based permutation test showed significant differences in the probability distributions of interactions for comparing ASM load conditions. These differences were expressed by an increased probability of fast next-interval dynamics following tapering, which reversed after restarting the medication (figure 2a).

**Figure 2.**
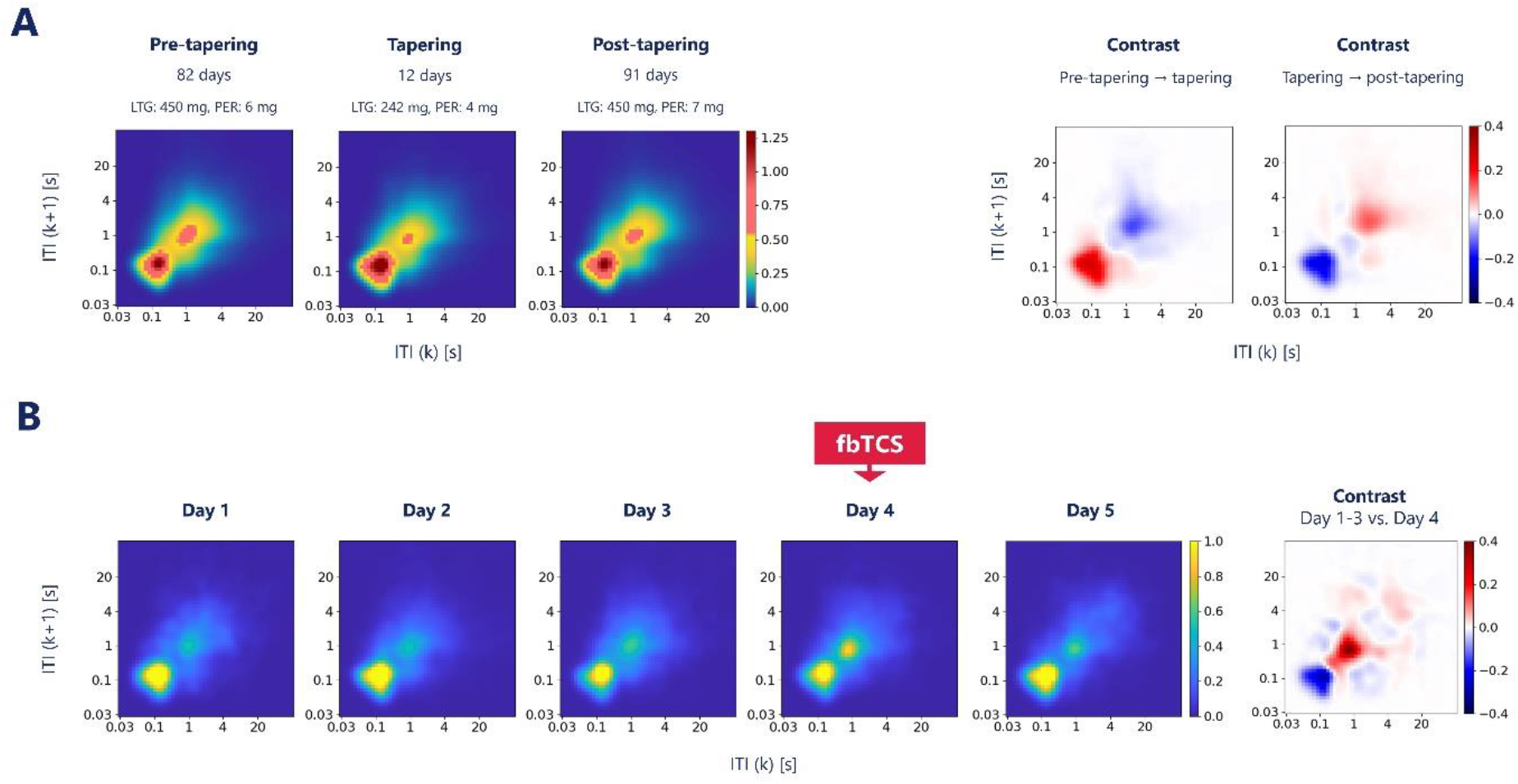
**A)** Comparison of touchscreen behavioural patterns between pre-tapering, tapering and post-tapering period. Left: daily joint interval distributions (JID) of inter-touch intervals averaged per period. Right: Contrasts between the daily JIDs from the pre-tapering and tapering periods and between the daily JIDs from the tapering and post-tapering periods. Cluster-based permutation testing revealed significant differences for both transitions (i.e. pre-tapering → tapering and tapering → post-tapering), reflected by an acceleration of the next-interval dynamics during tapering that reversed on medication restart. **B)** Comparison of touchscreen behavioural patterns between a day with a convulsion during 5-day admission. Shortly after rise time on day 4, the man experienced a convulsive seizure. The first five figures are JIDs of smartphone touchscreen interactions during admission. Far-right figure: the contrast between the JID of the day with fbTCS and the JIDs of the three days before. It shows a deceleration of the next-interval dynamics on the day of the fbTCS. **Abbreviations:** fbTCS: focal to bilateral tonic clonic seizure; ITI: inter-touch interval; LTG: lamotrigine; PER: perampanel

The probability distributions of the interactions showed marked contrasts between the days with and without focal to bilateral tonic-clonic convulsion, expressing an increased probability of slow next-interval dynamics on the day of the convulsion compared to the three preceding days (figure 2b).

## Discussion

We found marked behavioural changes following an focal to bilateral tonic-clonic convulsion and in response to ASM changes. These changes were recorded with a regular smartphone with a negligible additional burden to the user. Touchscreen interactions show an increased probability of slow next-interval dynamics following a convulsionand increasing ASM load. In contrast, tapering showed the opposite (i.e. an increased likelihood of fast next-interval dynamics). The pattern is similar to what is seen with increasing chronological age.^5,10^

ASMs users commonly report cognitive side effects^11^, and they are a robust predictor of reduced quality of life.^1^ Information on these effects is helpful during the titration of ASMs, but an adequate, unobtrusive tracking method is lacking. Seizures may also aggravate cognitive impairments or mood disorders,^12^ but are similarly challenging to monitor as self-reported seizure diaries are often inaccurate.^13^

This case suggests smartphones may address the need to track medication and seizure-related behavioural changes. It also underscores the complexity of the fluctuations in subject status that cannot be captured by cross-sectional assessments such as questionnaires or self-reports. The neurobehavioural substrates are still largely unexplored, but there is some evidence that touchscreen interactions correlate with conventional assessments of reaction time^14^ and reliably estimate biological age.^5^ Interestingly, epileptiform discharges coincide with distinct profiles, suggesting that even subtle changes can be monitored.^8^

We propose that the increased behavioural ageing in epilepsy is partly explained by ASM exposure and seizure occurrence. ASMs may cause a broad spectrum of cognitive changes, including impaired executive function, attention and reaction time.^15,16^ Our finding of significant behavioural changes following ASM load adjustment is important because the cognitive and behavioural side effects of the ASMs prescribed to the case (lamotrigine and perampanel) are relatively unknown but relevant.^17,18^ A more pronounced change in smartphone behaviour could be expected with ASMs associated with adverse cognitive effects or with polyteraphy.^15^ Notably, the net accelerating ASM effect was significant. However, the period also included the day of the convulsive event, which showed a reverse effect on smartphone behaviour (i.e. a deceleration of interactions).

We grouped touchscreen interactions into daily interaction profiles, as hourly profiles would yield variable amounts of data due to intermittent phone use. While the interaction profile of the day with focal to bilateral tonic-clonic convulsion included a pre- and post-ictal phase, it predominantly reflects the post-ictal phase as the seizure occurred soon after wake-up. The post-ictal state can manifest in a variety of ways, including altered consciousness and cognitive dysfunction.^19^ Cognitive impairments may last for hours or even days after a focal to bilateral tonic-clonic convulsion. Our case’s loss of rapid touchscreen interactions may well reflect these post-ictal signs.

We only examined the effects of the convulsion without evaluating the days with focal seizures. We anticipated that the cognitive impact of focal seizures, particularly those with retained awareness, is considerably less. Many seizures recorded and a sufficiently long seizure-free period are necessary to evaluate their effect on smartphone behaviour. We selected a single case to demonstrate the impact of medication withdrawal and the recorded convulsion. Cohort studies are needed to establish our findings’ generalisability and ascertain the potential of touchscreen profiles in epilepsy care.

## Data Availability

The data supporting this study's findings are available from the corresponding author upon reasonable request.

## Acknowledgements

This study was funded by Health-Holland, Top Sector Life Sciences & Health Netherlands Organization for Health Research and Development (ZonMW) [Brain@home, Project number: 114025101].

R.D.T. is supported by the Christelijke Vereniging voor de Verpleging van Lijders aan Epilepsie, and by research .grants from the Dutch National Epilepsy Fund (EpilepsieNL),

This study leveraged the support to A.G. by a research grant from Velux Stiftung (Project number: 1283).

## References

1. Perucca P, Carter J, Vahle V, Gilliam FG. Adverse antiepileptic drug effects: Toward a clinically and neurobiologically relevant taxonomy. Neurology. 2009; 72(14):1223–9.

2. Hermann BP, Struck AF, Busch RM, Reyes A, Kaestner E, McDonald CR. Neurobehavioural comorbidities of epilepsy: towards a network-based precision taxonomy. Nat Rev Neurol. 2021; 17(12):731–46.

3. Wan S, Liang Y, Zhang Y, Guizani M. Deep Multi-Layer Perceptron Classifier for Behavior Analysis to Estimate Parkinson’s Disease Severity Using Smartphones. IEEE Access. 2018; 6:36825–33.

4. Apolinário-Hagen J, Menzel M, Hennemann S, Salewski C. Acceptance of Mobile Health Apps for Disease Management Among People With Multiple Sclerosis: Web-Based Survey Study. JMIR Form Res. 2018; 2(2):e11977.

5. Ceolini E, Brunner I, Bunschoten J, Majoie MHJM, Thijs RD, Ghosh A. A model of healthy aging based on smartphone interactions reveals advanced behavioral age in neurological disease. iScience. 2022; 25(8):104792.

6. Baecker L, Garcia-Dias R, Vieira S, Scarpazza C, Mechelli A. Machine learning for brain age prediction: Introduction to methods and clinical applications. eBioMedicine. 2021; 72:103600.

7. Borger JN, Huber R, Ghosh A. Capturing sleep–wake cycles by using day-to-day smartphone touchscreen interactions. Npj Digit Med. 2019; 2(1):1–8.

8. Duckrow RB, Ceolini E, Zaveri HP, Brooks C, Ghosh A. Artificial neural network trained on smartphone behavior can trace epileptiform activity in epilepsy. iScience. 2021; 24(6):102538.

9. Ceolini E, Ghosh A. Common multi-day rhythms in smartphone behavior. Npj Digit Med. 2023; 6(1):1–9.

10. Ceolini E, Kock R, Band GPH, Stoet G, Ghosh A. Temporal clusters of age-related behavioral alterations captured in smartphone touchscreen interactions. iScience. 2022; 25(8):104791.

11. Kennedy GM, Lhatoo SD. CNS adverse events associated with antiepileptic drugs. CNS Drugs. 2008; 22(9):739–60.

12. Novak A, Vizjak K, Rakusa M. Cognitive Impairment in People with Epilepsy. J Clin Med. 2022; 11(1):267.

13. Mielke H, Meissner S, Wagner K, Joos A, Schulze-Bonhage A. Which seizure elements do patients memorize? A comparison of history and seizure documentation. Epilepsia. 2020; 61(7):1365–75.

14. Balerna M, Ghosh A. The details of past actions on a smartphone touchscreen are reflected by intrinsic sensorimotor dynamics. Npj Digit Med. 2018; 1(1):1–5.

15. Witt J-A, Helmstaedter C. Monitoring the cognitive effects of antiepileptic pharmacotherapy--approaching the individual patient. Epilepsy Behav EB. 2013; 26(3):450–6.

16. van Schooneveld MMJ, van Erp N, Boshuisen K, Meekes J, Braun KPJ. Withdrawal of antiepileptic drugs improves psychomotor speed after childhood epilepsy surgery. Epilepsy Res. 2013; 107(1–2):200–3.

17. Aldenkamp AP, Baker G. A Systematic Review of the Effects of Lamotrigine on Cognitive Function and Quality of Life. Epilepsy Behav. 2001; 2(2):85–91.

18. Rugg-Gunn F. Adverse effects and safety profile of perampanel: a review of pooled data. Epilepsia. 2014; 55 Suppl 1:13–5.

19. Pottkämper JCM, Hofmeijer J, van Waarde JA, van Putten MJAM. The postictal state — What do we know? Epilepsia. 2020; 61(6):1045–61.

